# Assessment of indoor biological air quality at a mass gathering event in an unimproved exhibition facility during the COVID-19 pandemic using a novel air sampling technology

**DOI:** 10.1101/2022.02.19.22271227

**Authors:** Julian Gordon, Osama Abdullah, Rachel Reboulet, Kara Hanson, Christine Sadowski, Hunter Rennels, Steve Kuemmerle, Richard Tuttle, Kristen Solocinski, Brittany Knight, Jacob Wilkinson, Gavin Macgregor-Skinner

## Abstract

The objective was to evaluate the determination of biomarkers of air quality during a mass gathering event at a convention center using a novel air sampling device, AirAnswers®. This sampler has previously only been used in smaller locations. Here it was run at five crowded locations within the exhibit area for the four days duration of a trade show. The AirAnswers® device uses electro-kinetic flow to sample air at high rates and capture bio-aerosols on grounded electrodes in assayable form. Cartridges were removed from the devices and immediately conveyed to the Inspirotec facility in North Chicago, where assays were performed.

Biomarkers determined were for allergens and molds previously described for this system. Testing for a new marker, severe acute respiratory syndrome coronavirus 2 (SARS-CoV-2) RNA was also included. The method was validated by determination of capture efficiency with reference to an impinger sampler in a Class III controlled environment chamber. Average capture efficiency for triplicate runs was 14%. One SARS-CoV-2 positive sample as found at the registration area, which was physically separate from the main exhibit area.

Cat allergen Fel d 1was found in four of the locations, dog allergen Can f 1 at two. The airborne biomarker of mold proliferation, (1→3)-β-D-Glucan, was above the assay range in all locations. The widespread presence of this mold marker could be accounted for by signs of water leakage. A generic 18S RNA marker for mold was developed and similarly showed the presence of mold in all locations, as was a genus marker for *penicillium*. A species marker for *Cladosporium cladosporioides* was in two locations. Species markers for *Eurotium amstelodami* and *Trichoderma viride* were each in a single location.

The main findings were of the widespread presence of mold markers, and the sporadic appearance of SARS-CoV-2. Masking was recommended but not enforced.

## INTRODUCTION

The AirAnswers® device for air sampling uses electro-kinetic flow to sample air at high rates, and capture bio-aerosols on the grounded electrodes in assayable form. Published data to date using this system [1-4] has been for single home locations and in various locations in a school system [5]. Previous measurements have focused on common household allergens and (1→3)-β-D-Glucan, a fungal cell-wall fragment. In this study, the utility is extended to the validation of the measurement of airborne SARS-CoV-2.

As scientific understanding of the routes of severe acute respiratory syndrome coronavirus 2 (SARS-CoV-2) the causative agent of Coronavirus disease 2019 (COVID-19) transmission progressed, experts in the field concluded that the main means of transmission was via fine droplets in the air [6]. This was based on identification of spreader events from the origin in Hunan, China and subsequent global spread. However, direct measurements of the airborne coronavirus have been limited. Heneghan et al placed online a comprehensive review of work on airborne SARS-CoV-2 and showed the need for a standardized validated procedure [7]. All work thus far has been based on limited numbers of observations and no formal validation of methods.

Methods that have been applied to attempts to detect airborne SARS-CoV-2 include glass fiber filter [8], gelatin filters [9] various other kinds of filters [10], condensate capture [11-14], NIOSH vortex sampler[15-17] electrostatic aerosol to hydrosol [18], SKC Impinger [19], personal environmental monitor [20], Sioutas Cascade Impactor [21].or bubbling through triazole [22]. One study made use of the AirAnswers® to show presence of airborne SARS-CoV-2 associated with the choir in a high school [5]. Here the measurement is validated in a controlled environmental chamber.

After being canceled in 2020, amid the COVID-19 pandemic, the trade show returned in 2021, and AirAnswers® devices were placed at five locations within the exhibit area. The facility had been unoccupied for two years, and only one other convention had been held immediately preceding this one. We analyzed air samples for biological agents that included allergens, mold, and SARS-CoV-2 virus.

## METHODS

### 1. Description of venue

At the request of the trade show organizers, identifiers to the actual congress have been kept anonymous. Air samples were taken at the positions indicated as 1-5 in Fig 1. The location was in a ballroom in a conference center during a four-day trade show for environmental remediation and cleaning professionals. The ballroom is 85,204 sq. ft, 358 ft width, 238 ft length and 30–34 ft high. The foyer was separated from the ballroom by full height walls and is 15,670 sq. ft. Positions of doorways into the ballroom area are indicated in Fig. 1.

**Fig. 1.**
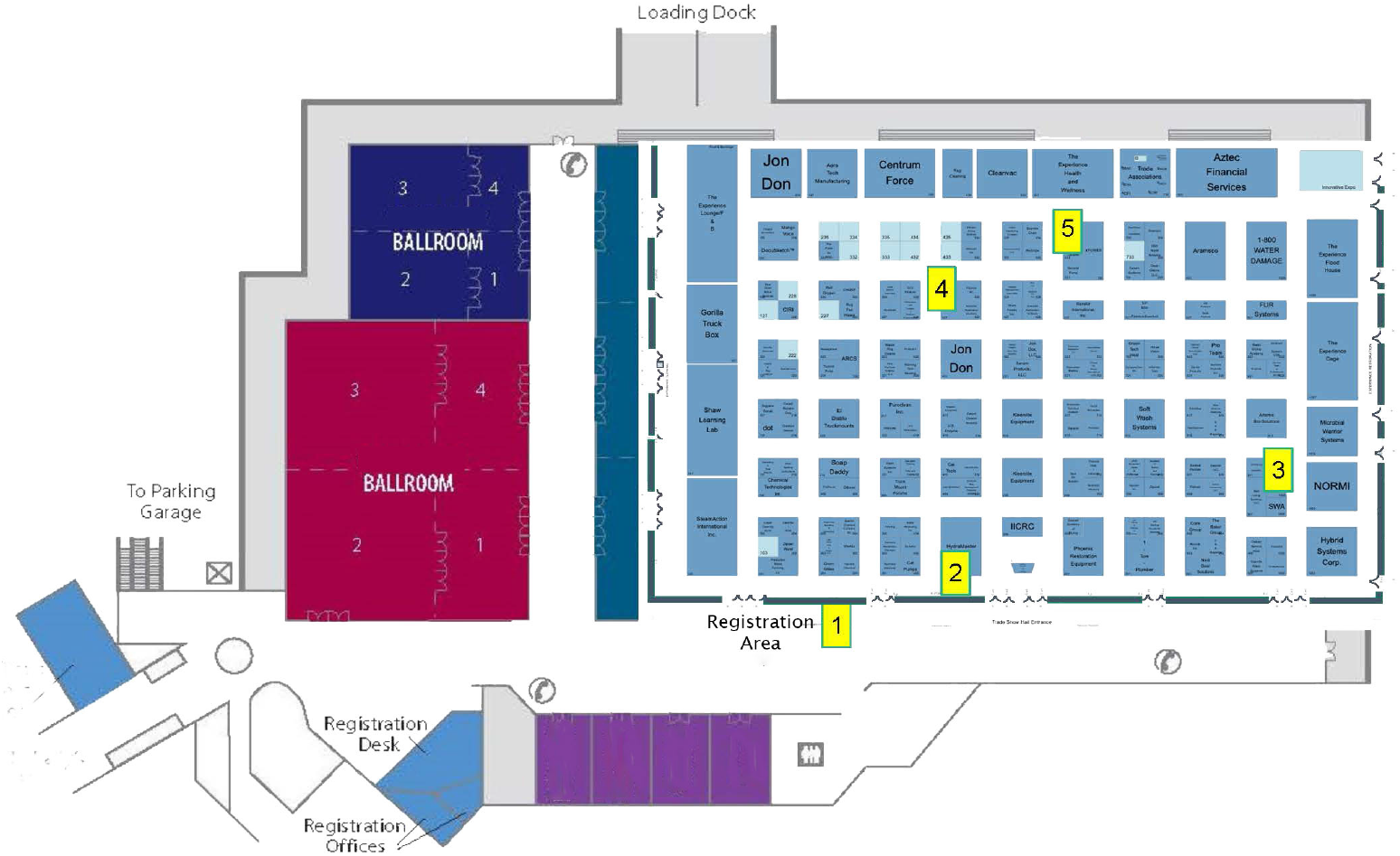
Layout of tradeshow. Exhibit area and locations of individual exhibits are shown. Dispositions of samplers are numbered locations in yellow boxes. Registration area is in a foyer outside of general exhibit area.

There were approximately 3500 people registered to attend the meeting. Of these 3500, it is estimated that up to 2500 people may have been in the ballroom at any one time. Masking or distancing were not strictly observed, but evidence of vaccination had been required of participants. Locations in high traffic and/or congregation areas, e.g. meeting registration areas, concourses, waiting areas, water stations, etc. were identified as possible positions for Air Answers™ device placements. Actual positions selected were in the registration area (1), at a water station where bottled water was handed out (2), AirAnswers® Booth (3), and other vendors’ booths at (4) (5). Multiple 30-minute seminars were conducted each day at booth (5). Permission for the placements were provided by the trade show organizers and no complaints were received about the presence of AirAnswers® devices.

AirAnswers® devices were run for four days for the duration of the trade show. Cartridges containing captured bioaerosol sample were removed from the devices and immediately conveyed to the Inspirotec facility in North Chicago where assays were performed.

### 2. Test methods

Multiplex immunoassays for allergens were performed at Indoor Biotechnologies (Charlottesille, Va) using their proprietary MARIA® technology for: pollens: Birch (Amb a 1), Ragweed (Bet v 1), Timothy grass (Phl p 5); Pets: Dog (Can f1), Cat (Fel d 1); Pests: Cockroach (Bla g 2) and Mouse (Mus m1). Assay of (1→3)-β-D-glucan, a measure of aerosolized fungal cell wall fragments, was performed using the kinetic Limulus Amebocyte assay (Associates of Cape Cod, Inc., Woods Hole, Massachusetts).

Species-specific mold spore analysis was on the insoluble fraction from the electrode extracts with the fungal-specific quantitative polymerase chain reaction (qPCR) for 23 species, provided by Eurofins (Eurofins EMLab P&K Chicago, Naperville, IL) based on EPA designs [23, 24]. Further, we have designed a set of genus-specific primers and probes based on the sequences listed by the EPA [25], see supplementary data, Table S1. A BLAST search was conducted on each set of primers and probes to confirm *in silico* specificity. In addition, total fungal load was measured with primers for a conserved region of the 18S RNA gene [25] (supplementary data Table S1)..

### 3. Validation of SARS-CoV-2 virus capture efficiency

Testing was performed at MRI-Global, an independent research organization, in a 2.5 ft X 3.5 ft 1.5 ft plexiglass Class III biosafety cabinet, total volume about 371 L. USA-WA1/2020 strain of virus (BEI Resources, Manassas, VA) was cultivated in Dulbecco Modified Eagles Medium with 5% fetal bovine serum, and penicillin, streptomycin and neomycin. Freshly grown stock at 3.16×10^6^ TCID50/mL was injected into the chamber. Test runs were for 30 minutes. The chamber contained triplicate AirAnswers® devices and a Midget Sample Impinger (Ace Glass) as reference method. Devices were placed in the chamber equidistant from the walls with a fan running for 10 minutes prior to run. A collision 6 Jet Nebulizer for aerosol generation, driven by HEPA filtered air, was filled with 10 mL of viral suspension prior to the run. An APS 3221 aerosol particle sizer pulled air for 30 seconds at times 0, 10 and 20 minutes, with dynamic particle size range 0.3 to 20 μm. Particle count results are given in Supplementary Data Table S2 for the three runs

The samples collected with the AirAnswers® device were assayed by real-time PCR with a BioRad CFX96 Thermocycler. A modified CDC guideline protocol was adjusted for air samples [26]. RNA Isolation was performed with (QIAGEN Viral RNA Mini Kit). N1 primers and probe, MasterMix and Positive control were provided by Integrated DNA Technologies, Thermo Fisher Scientific, and ATCC.

## RESULTS

### (1) Allergen at 5 locations in the Convention Center

AirAnswers® devices were run at 5 locations during the course of the trade show. Of the common airborne allergens tested, cat (Fel d 1) and dog (Can f 1) were detected (Table 2). Both were present in the registration area and Fel d 1 only was present in three booth locations. Can f 1 was additionally found at an area where multiple 30-minute seminars were conducted each day. The cat allergen is well known to be easily spread and could be carried on participants’ clothes. The fact that not all locations in the hall had the same allergen profile shows that there is not perfect mixing of the air during the four day course of the sampling.

**Table 1.** Capture efficiency of COVID-19 amplicons compared to impinger reference device.

| Sampler | Copies/mL | Copies/extr. | Flow rate(lpm) | Volume sampled L/run | Copies/L | Capture efficiency (%) |
| --- | --- | --- | --- | --- | --- | --- |
| 1 | 4.30E+06 | 8.60E+07 | 110 | 3300 | 2.61E+04 | 16.1 |
| 2 | 4.30E+06 | 8.60E+07 | 104 | 3120 | 2.76E+04 | 17.0 |
| 3 | 2.50E+06 | 5.00E+07 | 113 | 3390 | 1.47E+04 | 9.1 |
| Impinger | 6.80.E+05 | 6.80.E+06 | 1.4 | 42 | 1.62E+05 |  |

Copies/mL were from samples extracted with 10 mL DMEM per electrode and 10 mL DMEM initially in Impinger. PCR analysis of the extracts was as follows. The PCR plates included a five-point standard curve prepared using the synthetic quantitative SARS-CoV-2 RNA from ATCC (Cat # VR-3276SD) containing the ORF, E, and N genes at 10,000 copies/ $\mu\text{L}$ down to 1 copy/ $\mu\text{L}$ . All PCR reactions were performed in triplicate for each sample. The amplification within the range of the standard curve was verified for the Test Device collection plate sample extracts, impinger reference samples, and virus stock dilutions. The slope and y-intercept from the standard curve were then used to calculate the copies/mL of SARS- CoV-2 nucleocapsid RNA detected in the test samples, as well as the concentration of the stock virus. Number of amplicons per unit volume of air was calculated from the amount captured the volume of air sampled for each device. The capture efficiencies were determined with the reference method defined as 100%. The average capture efficiency for COVID-19 by the AirAnswers™ device was 14.1% (Table 1). At this value, the lower limit of detection (LLOD) for four days of sampling is approximately 1 copy per 5 M<sup>3</sup> of air.

**Table 2.** Allergen levels in air at 5 locations in conference center.

| Location | 1 | 2 | 3 | 4 | 5 |
| --- | --- | --- | --- | --- | --- |
| Allergen | Registration | Water station | Air Answers Booth | Booth | Seminar Booth |
| Amb a 1 | <0.09 | <0.09 | <0.09 | <0.09 | <0.09 |
| Bet v 1 | <0.3 | <0.3 | <0.3 | <0.3 | <0.3 |
| Bla g 2 | <1.7 | <1.7 | <1.7 | <1.7 | <1.7 |
| Can f 1 | **0.18** | <0.10 | <0.10 | <0.10 | **0.18** |
| Fel d 1 | **0.09** | <0.03 | **0.07** | **0.19** | **0.09** |
| Mus m 1 | <0.02 | <0.02 | <0.02 | <0.02 | <0.02 |
| Phl p 5 | <0.3 | <0.3 | <0.3 | <0.3 | <0.3 |
| (1→3)-β-D-Glucan | >100 | >100 | >100 | >100 | >100 |

### (2) Fungal presence in the air of Convention Center

All (1→3)-β-D-Glucan results were elevated above the measuring range of the assay (>100 pg/M^3^: Table 2). This suggests the extensive presence of mold or residues of mold in the building. Some water leak was noted in the ceiling above the water station (Location 2), but high values of (1→3)-β-D-Glucan were found in all areas. Therefore, more detailed analysis of mold speciation was undertaken making use of the availability of standard primer and probe sets as used for detecting species for calculation of relative moldiness index (RMI), as provided by Fintech. Table 3 lists the results for each species but we did not calculate RMI as RMI is based on values from spores collected in dust, and this study focused on measurement of airborne spores. The results are informative of the degree to which species detected, if any, will spread through the entire space. *Cladosporium cladosporioides* and *Trichoderma viride* are present in the two locations, the registration desk area and the AirAnswers® Booth (location 3 in Fig 1). This parallels the distribution of the dog allergen Can f 1. The Registration desk area is located in the foyer, which is walled off from the main exhibit, although there may be some air exchange through the connecting doors. The AirAnswers® Booth also shows the presence of Eurotium (Asp.) amstelodami, not detected in the other sample locations.

**Table 3.**
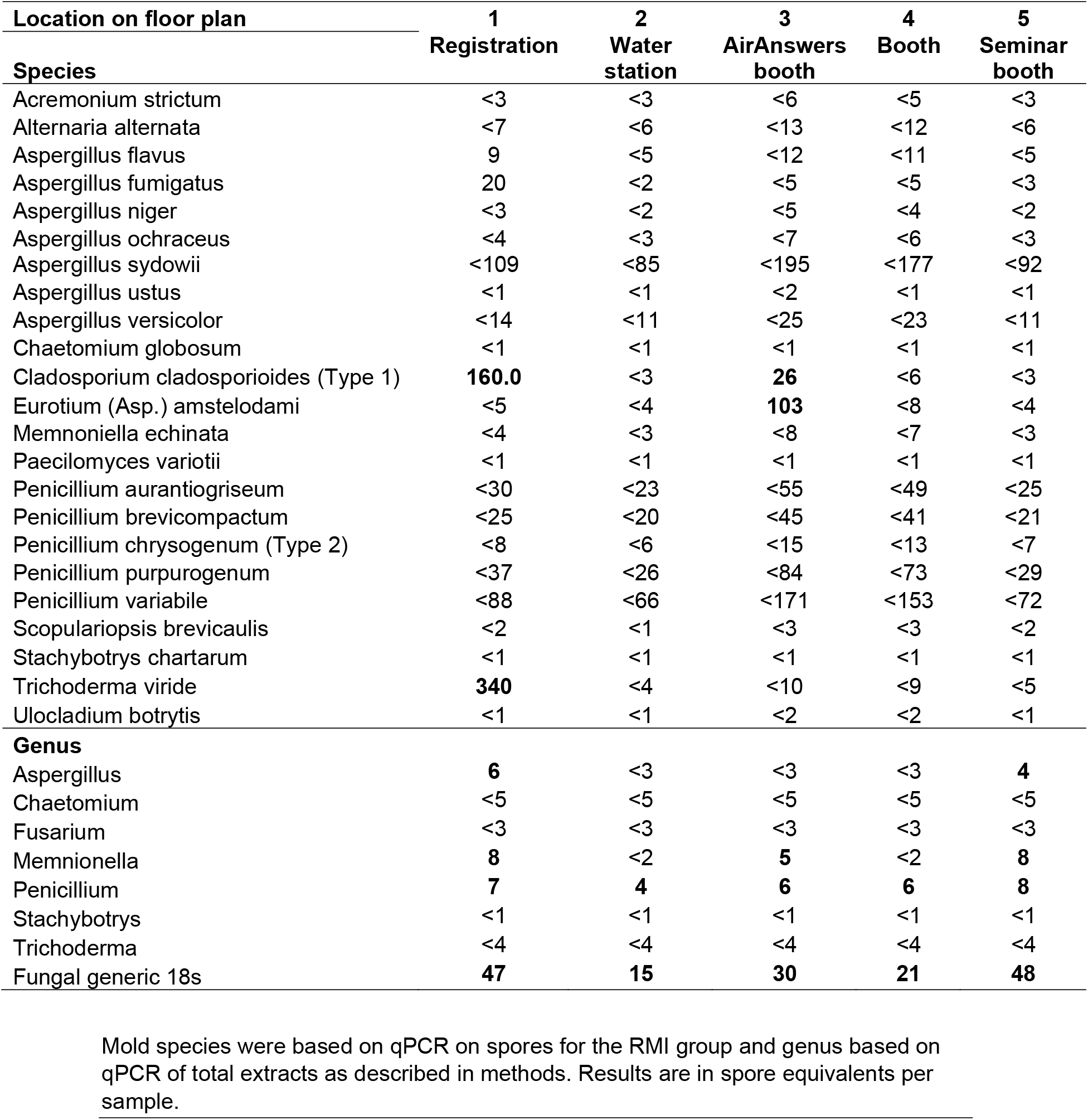
Mold genus and species at 5 location in the Convention Center

| Location on floor plan | 1 | 2 | 3 | 4 | 5 |
| --- | --- | --- | --- | --- | --- |
| Species | Registration | Water station | AirAnswers booth | Booth | Seminar booth |
| *Acremonium strictum* | <3 | <3 | <6 | <5 | <3 |
| *Alternaria alternata* | <7 | <6 | <13 | <12 | <6 |
| *Aspergillus flavus* | 9 | <5 | <12 | <11 | <5 |
| *Aspergillus fumigatus* | 20 | <2 | <5 | <5 | <3 |
| *Aspergillus niger* | <3 | <2 | <5 | <4 | <2 |
| *Aspergillus ochraceus* | <4 | <3 | <7 | <6 | <3 |
| *Aspergillus sydowii* | <109 | <85 | <195 | <177 | <92 |
| *Aspergillus ustus* | <1 | <1 | <2 | <1 | <1 |
| *Aspergillus versicolor* | <14 | <11 | <25 | <23 | <11 |
| *Chaetomium globosum* | <1 | <1 | <1 | <1 | <1 |
| *Cladosporium cladosporioides* (Type 1) | **160.0** | <3 | **26** | <6 | <3 |
| *Eurotium* (Asp.) *amstelodami* | <5 | <4 | **103** | <8 | <4 |
| *Memnoniella echinata* | <4 | <3 | <8 | <7 | <3 |
| *Paecilomyces variotii* | <1 | <1 | <1 | <1 | <1 |
| *Penicillium aurantiogriseum* | <30 | <23 | <55 | <49 | <25 |
| *Penicillium brevicompactum* | <25 | <20 | <45 | <41 | <21 |
| *Penicillium chrysogenum* (Type 2) | <8 | <6 | <15 | <13 | <7 |
| *Penicillium purpurogenum* | <37 | <26 | <84 | <73 | <29 |
| *Penicillium variabile* | <88 | <66 | <171 | <153 | <72 |
| *Scopulariopsis brevicaulis* | <2 | <1 | <3 | <3 | <2 |
| *Stachybotrys chartarum* | <1 | <1 | <1 | <1 | <1 |
| *Trichoderma viride* | **340** | <4 | <10 | <9 | <5 |
| *Ulocladium botrytis* | <1 | <1 | <2 | <2 | <1 |
| **Genus** |  |  |  |  |  |
| *Aspergillus* | **6** | <3 | <3 | <3 | **4** |
| *Chaetomium* | <5 | <5 | <5 | <5 | <5 |
| *Fusarium* | <3 | <3 | <3 | <3 | <3 |
| *Memnionella* | **8** | <2 | **5** | <2 | **8** |
| *Penicillium* | **7** | **4** | **6** | **6** | **8** |
| *Stachybotrys* | <1 | <1 | <1 | <1 | <1 |
| *Trichoderma* | <4 | <4 | <4 | <4 | <4 |
| Fungal generic 18s | **47** | **15** | **30** | **21** | **48** |

Mold species were based on qPCR on spores for the RMI group and genus based on qPCR of total extracts as described in methods. Results are in spore equivalents per sample.

The RMI primer-probe sets are well-established for mold species. Possible mold species may be missed by not being included in the RMI list. We, therefore, designed genus-specific probes and primers that may lead to results with more generality. The genus tests were done on total extract from the electrodes, so that, in addition to spores, any airborne fragments of fungus that included amplifiable DNA or DNA fragments would be included, if present. Results in Table 2 show Aspergillus was detected in the registration desk area, even though no species of aspergillus spores was found. Penicillium was detected at all 5 locations, even though no Penicillium spores had been detected at the species level. This suggests that some undefined Penicillium species spore or fragments was in the air of the entire facility, and that was not included in the RMI 23. Memnionella was detected at three locations of the facility, even though no Memnionella spores had been detected at the species level. Not unexpectedly, the fungal-generic 18S primer and probe set confirmed the presence of fungal material in the air throughout.

### (3). Presence of SARS-CoV-2 in the Exhibit Hall

SARS-CoV-2 was detected at the registration desk area and not at the other four sampling sites. This is an area of daily high-density crowding and high people traffic between the foyer and the main exhibit hall. The sampler at the registration desk area was positive at approximately 260,000 gene copies/M^3^ while four other sites were all negative at <1 gene copy/5 M^3^ of air. The presence of the virus only in the registration desk area could be explained by an infected individual or an asymptomatic virus shedder who either did not go on to visit the exhibition area, or that the air volume in the exhibit was so large that the shed virus was diluted to undetectable levels. This supports results above that a particular mold species may be confined to a relatively limited air space like that around the registration desk. Nevertheless, the occurrence of virus in the air at any location means that it is premature to relax restrictions that are intended to prevent occurrence of spreader events.

## DISCUSSION

With the advent of widespread vaccination and combined with masking and distancing, the risk of spread of the virus may be less, but this should not promote a false sense of security where precautions are relaxed. Indeed, the occurrence of waves of infection can be seen as feedback oscillations, when the frequency of hospitalizations and deaths is seen to decrease, public officials relax precautions to minimize damage to the economy, followed by rise of infection rates. This is exacerbated by the positive selection for more aggressive strains such as the delta variant and now the Omicron variant. The results presented here show that break-through events may occur with current practices. While it is not known to what extent the SARS-CoV-2 was associated with active disease or resulted in the spread of active disease technology for environmental surveillance to determine whether environmental measures are effective, and, if not, what remedial measures might be applicable for future events. The measurements of airborne molds and allergens also provide supportive information on the performance of whatever air-cleaning system is in place. Of those mold species identified as being present in the Convention (Table 3), Trichoderma viride, one of the zero tolerance molds, was detected along with Cladosporium. Cladosporium, one of the most allergenic molds, was detected in the registration desk area (1). Cladosporium was also located at the AirAnswers booth. For the air to be considered clean, the sources of these molds should be identified and eliminated.

It is noteworthy that no single strategy will eliminate the pandemic. Vaccination is never 100% effective. Masking and distancing lower the risk, but even masking does not completely eliminate the risk of infection. There is work suggesting that eyes may be a site for viral penetration [27] although eye protection is no longer widely used.

For a facility with an existing HVAC system, the cost of replacement or updating will depend on the specific facility. For a facility such as that which is the subject of this work, a more direct way of cleaning the air could be to add mobile air-cleaning devices throughout the facility. An example of a current system is that from Integrated Viral Protection portable air sterilization units (www.ivpair.com). Costs range from $750 to $9,999, depending on capacity. Their Venue Mobile Unit costs $13,995.00. This unit provides powerful circulation up to 30,000 cubic feet, over 3-5 changes an hour. This unit is built for large commercial spaces & facilities, delivering clean, virus-free indoor air [28]. The area of the Ballroom, which is the subject of the current work, is 85,204 sq ft. We estimate that 6 such units would be required to clean the air of this large space, at a cost of $84,000. This would be the minimal cost to remediate the air during a crowded congress, in the absence of a radical upgrade to the current HVAC system. In contrast, the analysis provided by AirAnswers would cost approximately $5,500. It is a cost-effective way of determining whether a more expensive remediation will be required. The presence of devices was accepted by organizers and no complaints received. A single measurement of SARS-CoV-2 above detection limits could be the initiation of a spreader event. The present study provides a baseline against which to monitor any future improvements of the air handling system. Karimzadeh et al [29] speculated that approximately 100 particles inhaled are sufficient to trigger an infection. From the concentration of particles measured at the registration desk, 260/L, an infectious dose would be inhaled in approximately 23 seconds, based on a standard breathing rate of 70 lpm. However, the PCR assays for SARS-CoV-2 may greatly overestimate the number of infectious particles in the air as non-viable damaged virus or viral fragments. There was no indication that this was a superspreader event as there was no documnted number of actual cases following the event. Spread may have also been mitigated by the organizer’s requirement that participants be vaccinated. Nevertheless, this provides a powerful motivation to ensure that indoor air quality should be monitored at mass gathering events especially during a pandemic.

## Data Availability

All data produced in the present work are contained in the manuscript

## ACKNOWLEDGEMENTS

This work was funded by Inspirotec Inc.

We gratefully acknowledge the help of Larry Cooper of The Experience for supporting the facilitation and the execution of this study

## SUPPLEMENTARY DATA

**Table S1:**
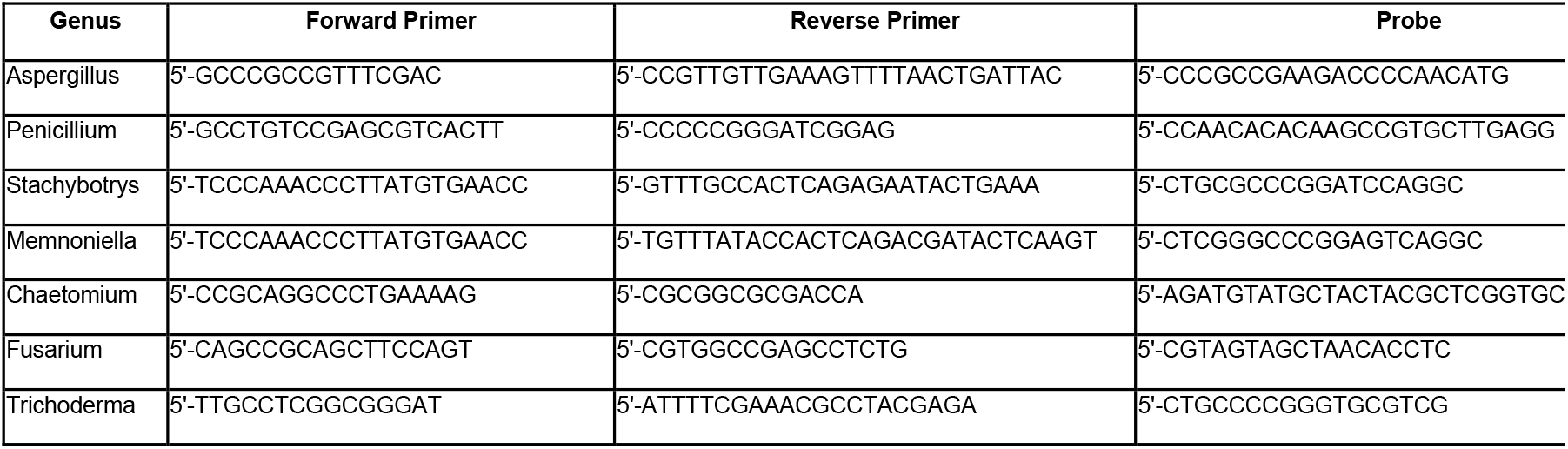
Primers and probes used for Genus-specific RT-PCR analysis’

**Table S2.** Particle levels during test runs of air samplers in environmental chamber.

| | Time<br>(min) | Particle<br>Counts | Mass<br>mg/m<sup>3</sup> | Median<br>diameter<br>(μm) |
| --- | --- | --- | --- | --- |
| Test 1 | 0 | 1,708,970 | 9.20 | 3.23 |
|  | 10 | 3,810 | 0.01 | 1.46 |
|  | 20 | 35 | 0.00 | 1.52 |
| Test 2 | 0 | 1,754,445 | 9.38 | 3.16 |
|  | 10 | 10,355 | 0.02 | 1.45 |
|  | 20 | 44 | 0.00 | 1.17 |
| Test 3 | 0 | 1,786,277 | 9.53 | 3.18 |
|  | 10 | 2,541 | 0.00 | 1.39 |
|  | 20 | 4 | 0.00 | 0.65 |

APS 3221 aerosol particle sizer measurements in environmental chamber in 3 independent runs taken for 10 seconds at times indicated. Note that although particles are rapidly depleted from the chamber due to capture by the three AirAnswers devices, they are subject to the same environment as the impinger reference sampling device.

## Notes

### Competing Interest Statement

JG, OA, RR, KH, CS, HR and SK are employees or contractors of Inspirotec Inc receiving remuneration, stocks or options and therefore have an interest in the commercial success of Inspirotec. RT, KS, BK and JW are employees of MRI-Global, performers of independent research with no interest in the outcome of the work. GM is an employee of the Global Biorisk Advisory Council, which performs accreditations and has no financial interest in the outcome of this work.

### Funding Statement

This study was funded by Inspirotec Inc

